# A major population resource of 474,074 participants in UK Biobank to investigate determinants and biomedical consequences of leukocyte telomere length

**DOI:** 10.1101/2021.03.18.21253457

**Authors:** V. Codd, M. Denniff, C. Swinfield, S.C. Warner, M. Papakonstantinou, S. Sheth, D.E. Nanus, C.A. Budgeon, C. Musicha, V. Bountziouka, Q. Wang, R. Bramley, E. Allara, S. Kaptoge, S. Stoma, T. Jiang, A.S. Butterworth, A.M. Wood, E. Di Angelantonio, J.R. Thompson, J.N. Danesh, C.P. Nelson, N. J. Samani

**Affiliations:** Department of Cardiovascular Sciences, University of Leicester, Leicester, United Kingdom; NIHR Leicester Biomedical Research Centre, Glenfield Hospital, Leicester, United Kingdom; School of Population and Global Health, University of Western Australia, Australia; British Heart Foundation Cardiovascular Epidemiology Unit, Department of Public Health and Primary Care, University of Cambridge, Cambridge, United Kingdom; National Institute for Health Research Blood and Transplant Research Unit in Donor Health and Genomics, University of Cambridge, Cambridge, United Kingdom; British Heart Foundation Centre of Research Excellence, University of Cambridge, Cambridge, United Kingdom; Health Data Research UK Cambridge, Wellcome Genome Campus and University of Cambridge, Cambridge, United Kingdom; Medical Research Council Biostatistics Unit, Cambridge Institute of Public Health, University of Cambridge, Cambridge, United Kingdom; The Alan Turing Institute, London, United Kingdom; Department of Health Sciences, University of Leicester, United Kingdom; Department of Human Genetics, Wellcome Sanger Institute, Hinxton, United Kingdom

## Abstract

The determinants and biomedical consequences of variation in leukocyte telomere length (LTL), a proposed marker of biological age, are only partially understood. Here we report the creation and initial characterization of LTL measurements in 474,074 participants in UK Biobank. We confirm that older age and male sex associate with shorter LTL, with women on average ∼7 years younger in “biological age” than men. Compared to white Europeans, LTL is longer in African, Chinese and other major ancestries. Older paternal age at birth is associated with longer individual LTL. Higher white cell count is associated with shorter LTL, but proportions of white cell subtypes have weaker associations. Age, ethnicity, sex and white cell count explain ∼5.5% of LTL variance. Using paired samples from 1351 participants taken ∼5 years apart, we show the regression-dilution ratio for LTL is ∼0.65. This novel resource provides major opportunities to investigate LTL and multiple biomedical phenotypes.

---

Many cardiovascular, neurodegenerative, neoplastic and other conditions increase in incidence with age. However, as suggested by substantial inter-individual variations in age of onset and disease risk^1^, these conditions are not inevitable consequences of aging. We and others have proposed that such variations may, at least in part, reflect variation in biological aging driven by variation in telomere length^2,3^. Telomeres are nucleoprotein complexes at chromosome ends that maintain genomic stability. They shorten with each cell division and determine cellular lifespan^4^. At a cellular level, mean telomere length (TL) reflects cellular age and replicative history^5^. Because of these and other properties, TL has been proposed as a biomarker of biological age^2^.

At a population level, TL has frequently been studied using leukocyte DNA, a practicable measure of TL that correlates well with TL across different tissues within individuals^6^. Leukocyte telomere length (LTL) shows considerable inter-individual variation and is largely genetically determined, with heritability estimates of ∼0.70 ^7^. Even so, established genetic risk factors explain only a small fraction of the variation in LTL^8,9^. Age, sex, paternal age at birth and ethnicity are associated with LTL, but also account only for a small proportion of the inter-individual variation in LTL^7,10-14^. Even after taking these factors into account, several biological, behavioural and environmental characteristics correlate with, and potentially modify, LTL, including oxidative stress, inflammation, obesity, smoking, physical activity and dietary intake^15-18^. It remains uncertain, however, whether they are correlates or causative determinants. Furthermore, there is uncertainty about LTL’s degree of within-individual variation over time^19,20^.

Congenital premature aging syndromes arise from extreme shortening of telomeres due to rare mutations in telomere regulatory genes^21^. By contrast, more subtle inter-individual variation in LTL has been linked to risks of several common disorders in middle- and later-life, including certain cancers, coronary artery disease, Alzheimer’s disease, osteoarthritis, and lung diseases^22-26^. For many reported LTL-disease associations, however, it remains uncertain whether they chiefly reflect cause-and-effect relationships. For some conditions (e.g., coronary artery disease) causality is supported by associations between genetically-determined variation in LTL and disease risk^8^. However, even when causality is likely, studies have been insufficiently powered to characterize dose-response relationships of LTL with new-onset (“incident”) disease outcomes, even though this is needed to define risk thresholds.

Population biobanks afford significant opportunities to address the key uncertainties outlined above. However, insight into the determinants and biomedical consequences of LTL has been limited by the inability of biobanks to *combine* key study attributes. In particular, studies require robust LTL measurement, long-term follow-up of participants for incident disease outcomes, and exceptional statistical power. Studies also need detailed genomic information on participants, both to characterize the genetic architecture of LTL and to derive genetic “instruments” to enable Mendelian randomization analyses to help judge causality. Importantly, studies also require extensive biomedical phenotyping, including information on behaviours, physiological traits and clinically relevant endpoints. Finally, studies require serial measurements, at least in subsets of participants, to enable quantification and correction for within-individual variation in LTL (“regression-dilution”) over time^27^.

UK Biobank (UKB) is a large population cohort established between 2006 and 2010 of participants aged 40-69 years at recruitment^28^. Participants have been characterised in detail using questionnaires, physical measurements, urinary and plasma biomarker measurements, genomic assays and longitudinal linkage with multiple health record systems^29^. Detailed imaging assessments of the brain, neck, heart, abdomen, bones and joints, and eyes have been conducted in large subsets of participants, as well as repeat blood sampling in several thousands of participants. Here, we report on the creation, quality assurance, and initial interrogation of a resource of LTL measurements in DNA samples of 474,074 participants in UKB. Our analyses highlight the scope and potential of this powerful and detailed resource, which is available to the worldwide research community through application to UKB.

## Results

### LTL measurements in 488,400 participants

Of the 489,090 DNA samples received by our laboratory from UKB, 488,400 remained after removal of duplicates and samples from participants who had withdrawn from the study (**Methods**; **Figure 1**). Valid LTL measurements were obtained for 474,074 (97.1%) samples. Of the 14,326 (2.9%) participants without a valid LTL measurement, the large majority had insufficient DNA, with only 1647 repeatedly failing LTL assay QC (**Figure 1**). A small proportion of participants had LTL measured in DNA samples not collected at baseline (**Figure 1**).

**Figure 1.**
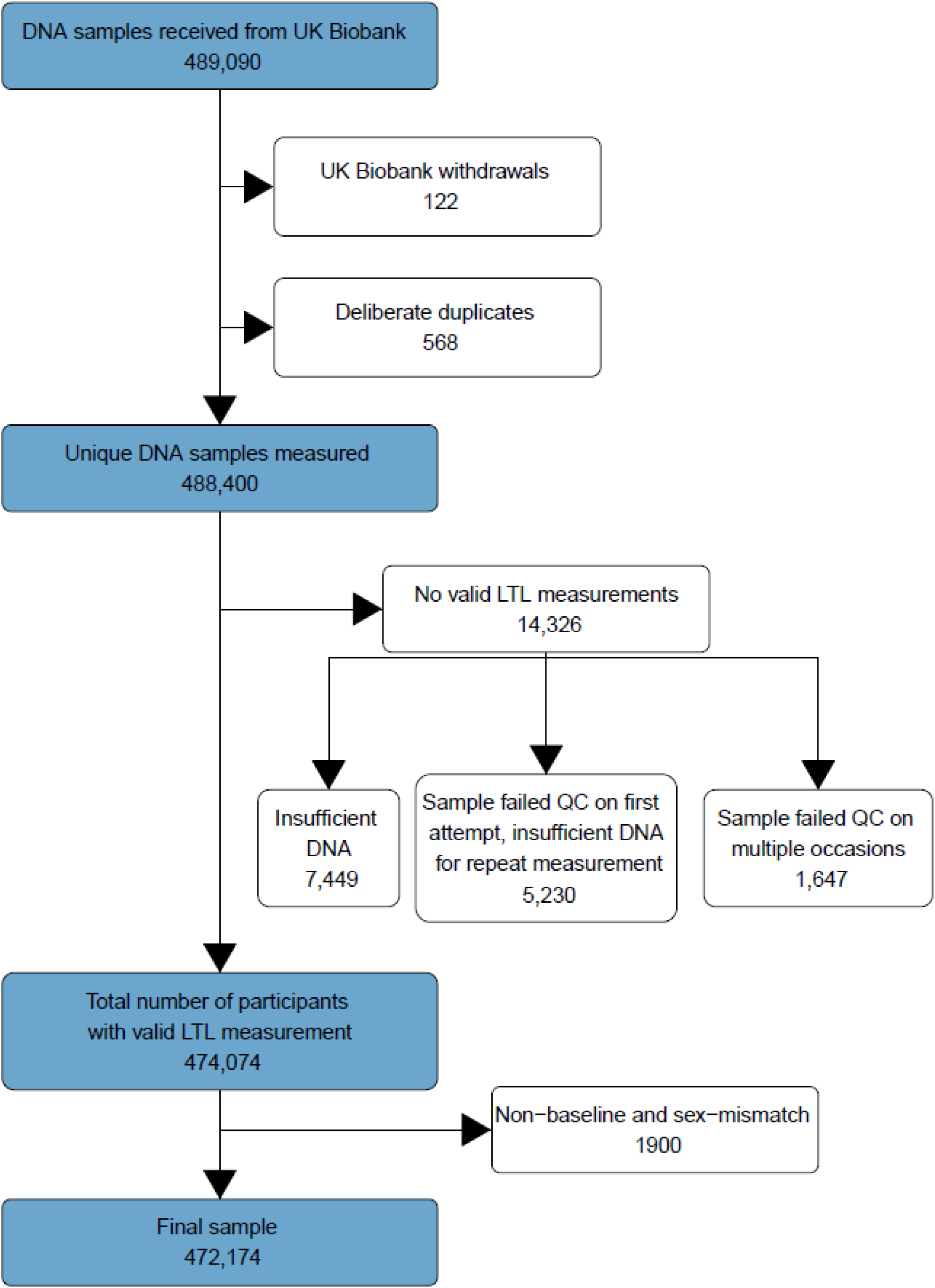
DNA sample workflow to derive the final dataset. After removal of study withdrawals and deliberate duplicate samples there were 488,400 participants for whom we attempted to measure LTL. Either a valid measurement was obtained or the sample was attributed to one of three categories of failure. For the downstream analyses presented in this paper that related to baseline phenotypes, we removed 1,900 DNA samples whose LTL was measured in a non-baseline DNA sample or where self-reported sex and genetic sex did not match.

Using multivariable regression models, we assessed the contribution of nine technical parameters to LTL variability, of which six had significant associations (**Table 1** and **Supplementary Figure 1**). PCR machine (Rotor-Gene Q), explained the greatest proportion of LTL variation in the multivariable model, followed by enzyme batch, temperature, staff member (operator), primer batch, and humidity. No associations were observed for the time of day of assay runs, pipetting robot (Qiagility) or DNA extraction method. We then considered all possible pairwise interactions and identified statistically significant interactions of primer batch with each of operator and PCR machine (**Table 1** and **Supplementary Figure 2**). In combination, the significant technical parameters and interactions explained 23.7% of LTL variation. Further, we estimated that the A260/280 ratio (a measure of DNA purity) explained an additional 0.5% of LTL variation (**Supplementary Figure 3**).

**Table 1.** Estimating the variance explained by each technical parameter. Data during stage 1 and 2 were assessed at the run level with linear regression on mean LTL. Stage 1: Univariate model R^2^ includes only this variable, multivariable partial R^2^ is the contribution of the parameter on the total model R^2^ (estimated as the difference between the full model R^2^ and the model R^2^ leaving this parameter out). Stage 2: Estimating the variance explained by the interactions in addition to the full model selected during Stage 1. Stage 2 Model R^2^=23.7%.

| Technical parameter | Univariate<br>Model R<sup>2</sup> (%) | Multivariable<br>partial R<sup>2</sup> (%) |
| --- | --- | --- |
| Stage 1 |  |  |
| Enzyme | 7.87 | 4.63 |
| PCR machine | 6.69 | 7.43 |
| Primer | 4.87 | 1.04 |
| Operator | 2.28 | 2.51 |
| Temperature | 0.73 | 4.63 |
| Humidity | 0.10 | 0.07 |
| Hours from 6am | 0.03 | - |
| Pipetting robot | 0.01 | - |
| Extraction method | 0.00 | - |
| Stage 2 |  |  |
| Primer* PCR machine | - | 2.13 |
| Primer*Operator | - | 1.56 |

To assess the impact of adjusting LTL for the relevant technical parameter mentioned above, we considered the mean LTL per week over the four-year assay period (**Figure 2**). While the unadjusted LTL measurements showed substantial fluctuations over time (**Figure 2A**), the adjusted LTL measurements were much more consistent across the assay period (**Figure 2B**). Adjustment strengthened the inverse correlation of LTL with age from −0.185 to −0.195 and increased the variance in LTL explained by age and sex from 4.04% to 4.53% (see further analyses below). Except when stated otherwise, the remaining analyses in this report used LTL values adjusted for technical parameters.

**Figure 2.**
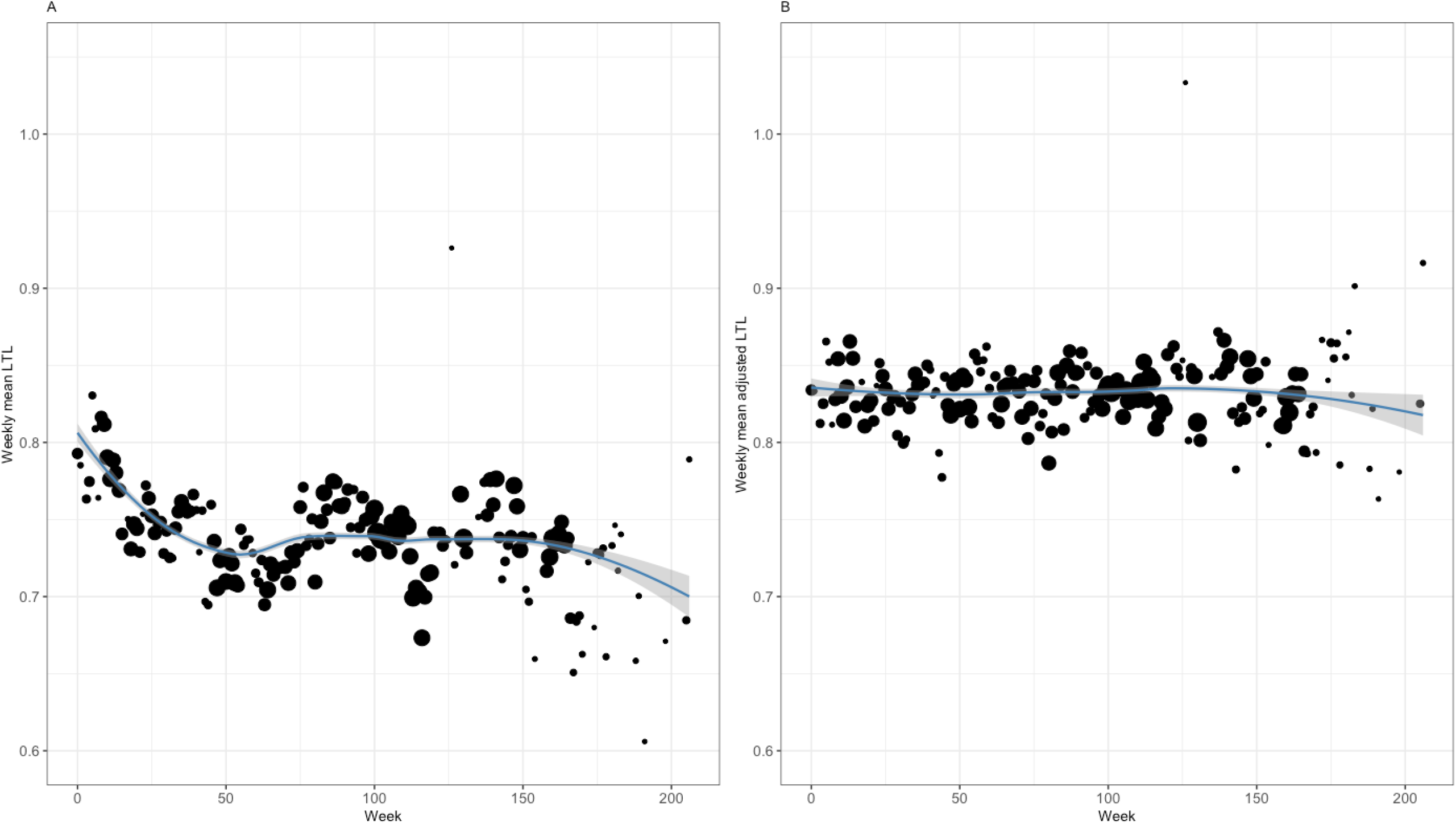
Distribution of weekly average LTL across duration of the study. A) The unadjusted LTL trend over time (and 95% confidence band). B) The adjusted LTL trend over time (and 95% confidence band). Adjustments for enzyme, PCR machine, primer, Operator, temperature, humidity, primer*PCR machine, primer*Operator and A260/280 were made as described in **Methods**. The smoothed curve is based on half plate means, with plotted data points representing overall weekly means. The size of each point indicates the number of runs that week. There were fewer measurements made after week 175, reflecting the period that sample QC and re-measurements took precedence following QC checks towards the end of the project.

### Reproducibility of LTL measurements

To assess our assay’s reproducibility, we calculated the coefficient of variation (CV) using samples measured on two separate occasions. For the blinded duplicates (n=528) included by UKB, the distribution of CVs was strongly positively skewed (**Supplementary Figure 4A**), with median CVs of 7.15 (IQR 3.03-11.69) for the raw LTL measurements and 6.53 (IQR 2.87-11.30) for the adjusted LTL measurements. For a larger set of randomly selected but unblinded repeats (n=22,516), the distribution of CVs was similarly skewed (**Supplementary Figure 4B**) with median CVs of 5.23 (IQR 2.44-6.33) and 5.53 (IQR 2.67-9.68) for the raw and adjusted values respectively.

To quantify within-person variability of LTL values over time, we calculated the regression-dilution ratio (RDR; see **Methods**) using 1351 available serial measurements of LTL taken at a mean interval of 5.5 years (range: 2-10 years). The RDR for LTL was 0.65 (95% CI: 0.61, 0.68) − similar to that for log_e_-transformed LTL (0.68, 95% CI: 0.64, 0.72) − and did not change materially with increasing time between serial measurements or after adjustment for participants’ age at sample collection (**Supplementary Figures 5A & 5B**). The well-known correlations of LTL with age, sex and other factors among participants with serial LTL measurements were similar to those in the entire UKB cohort (below and **Supplementary Figure 6**).

### Relationship between LTL and selected phenotypes

For these analyses, we focused on participants with LTL measurements on samples collected at UKB’s baseline examination, to match the time when the selected phenotypes were assessed (**Figure 1**). We also removed individuals where self-reported sex and genetic sex did not match, leaving 472,174 participants for these analyses. Characteristics of these participants, stratified by quartile of LTL values, are shown in **Table 2**.

**Table 2.**
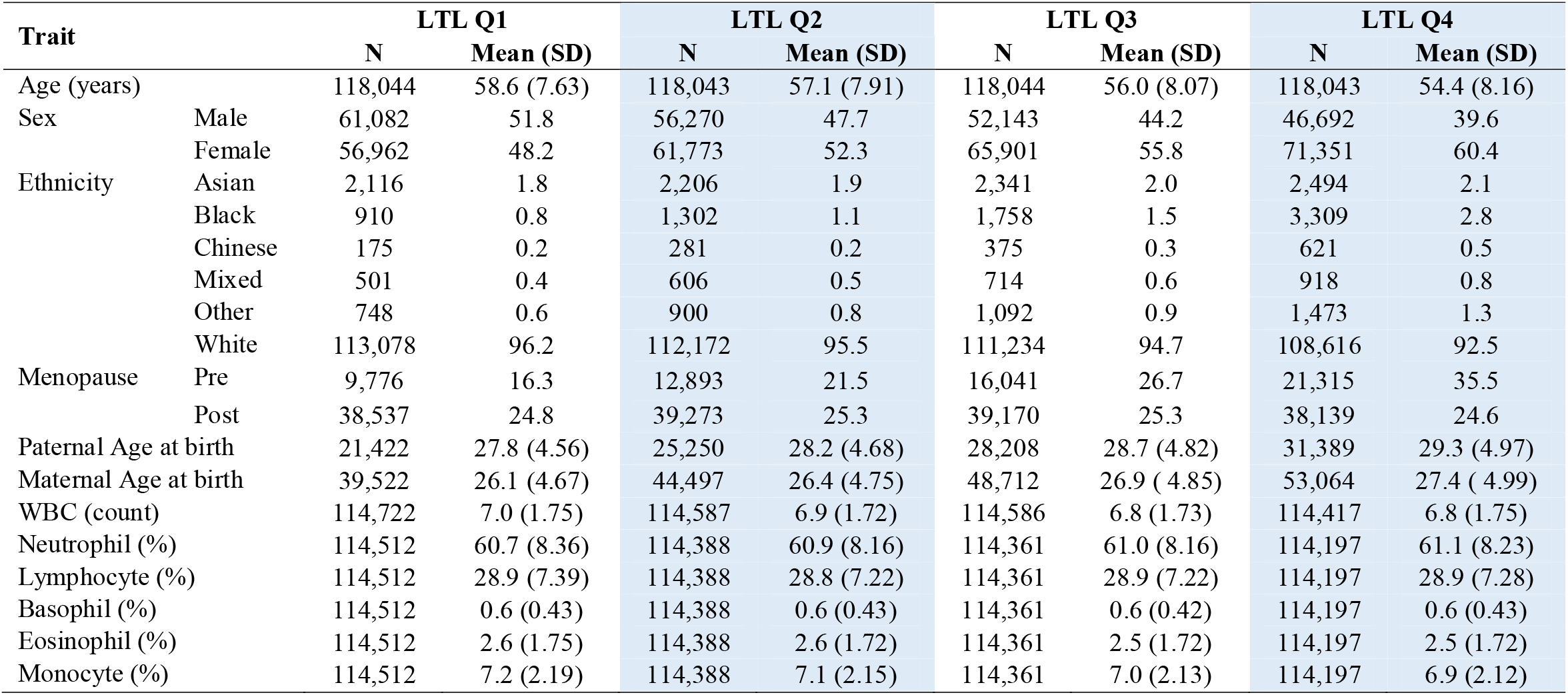
Characteristics of participants with LTL measurements at baseline. Data are shown by LTL quartile with Q1 being shortest LTL and Q4 being longest LTL. N is the available sample size, and the summary statistic is either the mean (standard deviation) for continuous traits or percentage for categorical traits. Ethnicity is self-reported and presented as defined by UKB Data-Field 21000. The Z-standardised values of LTL for each quartile are: Q1, <-0.65; Q2,-0.65≤ to <-0.002; Q3, −0.002 to <0.65; Q4, ≥0.65.

#### Age and sex relationships

We confirmed the known relationships between shorter LTL and older age and male sex (**Table 3** and **Supplementary Figure 7**). By comparing these associations, we estimated that women were on average about 7.4 years younger in “biological age” than men. Overall, the inverse association of LTL with older age was steeper in in men than women (**Table 3**; P=8.8×10^-37^ for age-sex interaction). Fitting a quadratic term for age within the model to men and women separately showed an almost linear inverse association among men of (P=0.034), compared to a shallower non-linear association in younger women that became steeper at older ages (P=3.80×10^-16^, **Supplementary Figure 8**). Further exploration showed that the steepness of the inverse association of LTL with age in women became similar to that in men after the menopause, particularly among post-menopausal women aged >55 years (**Table 3**).

**Table 3:** Relationship between LTL and age and sex. All models shown are fit with LTL as the outcome with available sample size N. *Model 1* includes age and sex. *Model 2* adds an interaction term between age and sex. *Models 3* (pre-menopausal), *4* (post-menopausal), 5 (aged ≤55 years) and 6 (aged>55 years) assesses age in sex stratified models where each woman is matched to a man of the same age before stratification. Betas are shown in SDs of LTL

| Model | N | Trait | Beta (95% CI) | P value |
| --- | --- | --- | --- | --- |
| **1** |  |  |  |  |
| Age and Sex | 437,544 | Age | -0.024 (-0.025, -0.024) | <1.00E-314 |
|  |  | Sex (Male) | -0.178 (-0.184, -0.172) | <1.00E-314 |
| **2** |  |  |  |  |
| Age and sex interaction | 437,544 | Age | -0.022 (-0.023, -0.022) | <1.00E-314 |
|  |  | Sex | 0.086 (0.045, 0.127) | 4.50E-05 |
|  |  | Age*Sex interaction | -0.005 (-0.005, -0.004) | 8.80E-37 |
| **3** |  |  |  |  |
| Pre-menopausal age matched | 54,560 | Male Age | -0.028 (-0.030, -0.026) | 2.00E-182 |
|  | 54,560 | Female Age | -0.023 (-0.024, -0.021) | 6.00E-116 |
| **4** |  |  |  |  |
| Post-menopausal age matched | 141,692 | Male Age | -0.027 (-0.028, -0.026) | <1.00E-314 |
|  | 141,692 | Female Age | -0.024 (-0.025, -0.023) | <1.00E-314 |
| **5** |  |  |  |  |
| Pre-menopausal aged ≤55 years age matched | 53,407 | Male Age | -0.027 (-0.029, -0.025) | 9.00E-122 |
|  | 53,407 | Female Age | -0.022 (-0.024, -0.020) | 7.20E-79 |
| **6** |  |  |  |  |
| Post-menopausal aged >55 years age matched | 111,962 | Male Age | -0.029 (-0.031, -0.028) | <1.00E-314 |
|  | 111,962 | Female Age | -0.029 (-0.030, -0.027) | 4.00E-302 |

#### Ethnicity

Compared to white Europeans, mean LTL was longer in people of Black, Chinese and mixed ancestries (**Supplementary Figure 9**). Within each ethnic group, we observed similar relationships of shorter LTL with older age and male sex (**Table 4**) to those reported overall, with somewhat steeper associations with age in Black participants (**Table 4**; **Supplementary Figure 10**). Differences in “biological age” between women and men across ethnic groups ranged from 5.88 years for South Asians and other Asians to 7.56 for Chinese.

**Table 4.** Age and sex associations within ethnic groups. A linear regression on LTL stratified by ethnicity and adjusting for age and sex. The age association is estimated for a single year increase in age adjusted for sex, and the sex association is the average difference in LTL for men compared to women adjusted for age. Ethnicity is self-reported and presented as defined by UKB Data-Field 21000. Betas are shown in SDs of LTL.

| Ethnic group | Age effect |  | Sex effect (Male) |  |
| --- | --- | --- | --- | --- |
|  | Beta (95% CI) | P value | Beta (95% CI) | P value |
| Asian | -0.026 (-0.028, -0.024) | 3.00E-95 | -0.153 (-0.195, -0.112) | 3.50E-13 |
| Black | -0.031 (-0.034, -0.028) | 6.70E-92 | -0.225 (-0.273, -0.177) | 8.40E-20 |
| Chinese | -0.025 (-0.032, -0.019) | 4.30E-14 | -0.189 (-0.292, -0.085) | 3.60E-04 |
| Mixed | -0.024 (-0.028, -0.019) | 1.30E-22 | -0.156 (-0.234, -0.077) | 1.10E-04 |
| Other ethnic group | -0.024 (-0.028, -0.020) | 1.60E-32 | -0.240 (-0.303, -0.178) | 7.00E-14 |
| White | -0.024 (-0.024, -0.023) | <1.0E-314 | -0.176 (-0.182, -0.170) | <1.0E-314 |

#### Paternal and maternal age at birth

Information on paternal and maternal age at birth was available for 97,234 and 170,668 participants, respectively, and on both parents for 70,871 participants. After adjustment for age and sex, having an older father or mother at birth was associated with longer LTL, including in analyses restricted to participants with information on both parents. The positive association per year of older parental age at birth with longer LTL was broadly equivalent to the *inverse* association per year of the participant’s age with shorter LTL (**Table 5**). Results were unchanged when restricting analyses for maternal (0.018, 95% CI: 0.016, 0.019) and paternal (0.021, 95% CI: 0.019, 0.022) age at birth only to participants with both parents alive at baseline. Including both maternal and paternal age at birth within the same model greatly attenuated the association of maternal age with LTL (**Table 5**), suggesting paternal age at birth is the principal determinant and that the relationship with maternal age at birth was likely due to correlation between parental ages (r=0.75), despite no evidence of collinearity (variance inflation factor [VIF]=2.29 and 2.26 for paternal and maternal ages, respectively). When we restricted analysis to participants with parental ages with a difference of between 2-5 years and >5 years, we found significant positive associations and consistent effect sizes with paternal age at birth but not with maternal age at birth (**Table 5**).

**Table 5.** The relationship between parental age at birth and LTL. Stage 1 analyses were performed in the whole cohort where the association with parental age was considered separately for paternal (n=97,234) and maternal (n=170,668), before fitting both in the regression model (n=70,871). Stage 2 analyses stratified by the age difference between both parents at birth to allow for the potential impact of the age difference driving the stronger paternal age association. Betas are shown in SDs of LTL.

| Stage 1 |  |  |  |  |  |  |
| --- | --- | --- | --- | --- | --- | --- |
| Trait | Paternal age only |  | Maternal age only |  | Parental age |  |
|  | Beta (95% CI) | P-value | Beta (95% CI) | P-value | Beta (95% CI) | P-value |
| Age | -0.022 (-0.023, -0.021) | 1.00E-314 | -0.023 (-0.024, -0.022) | 1.00E-314 | -0.022 (-0.024, -0.021) | 3.00E-270 |
| Sex | -0.139 (-0.151, -0.127) | 6.00E-111 | -0.151 (-0.160, -0.141) | 3.00E-223 | -0.130 (-0.145, -0.116) | 4.00E-72 |
| Paternal age | 0.020 (0.019, 0.022) | 5.00E-209 | - |  | 0.018 (0.015, 0.020) | 1.50E-52 |
| Maternal age | - |  | 0.017 (0.016, 0.018) | 1.00E-275 | 0.004 (0.002, 0.007) | 5.80E-04 |
| Stage 2 |  |  |  |  |  |  |
| Trait | 2 - 5 years |  | >5 years |  |  |  |
|  | Beta (95% CI) | P-value | Beta (95% CI) | P-value |  |  |
| Age | -0.021 (-0.023, -0.019) | 7.70E-75 | -0.022 (-0.026, -0.019) | 4.80E-36 |  |  |
| Sex | -0.120 (-0.146, -0.095) | 2.80E-20 | -0.118 (-0.154, -0.082) | 2.20E-10 |  |  |
| Paternal age | 0.018 (0.012, 0.024) | 3.70E-10 | 0.019 (0.014, 0.024) | 6.00E-15 |  |  |
| Maternal age | 0.004 (-0.001, 0.010) | 0.120 | 0.001 (-0.003, 0.006) | 0.550 |  |  |

#### White blood cells

In a model that also included age, sex and ethnicity, we found an inverse association of LTL with total white cell count (WBC) (0.064 SD lower LTL per 1-SD higher white cell count, p<1×10^-314^: **Table 6**). For individual white cell types, there was a positive association of LTL with proportion of neutrophils and inverse associations with proportions of eosinophil and monocytes. There was no association with lymphocyte percentage (**Table 6**).

**Table 6.** Multivariable model on LTL. Partial R^2^ is the contribution of the parameter on the total model R^2^ (estimated as the difference between the full model R^2^ and the model R^2^ leaving this parameter out). Total model R^2^ is 5.52%. Betas are shown in SDs of LTL.

| **Trait** | **Beta** | **P value** | **Partial R<sup>2</sup>** |
| --- | --- | --- | --- |
| Age (years) | -0.023 (-0.024 , -0.023) | <1.0E-314 | 3.52% |
| Male (ref: Female) | -0.170 (-0.176 , -0.164) | <1.0E-314 | 0.68% |
| WBC | -0.064 (-0.067 , -0.061) | <1.0E-314 | 0.37% |
| Neutrophil percentage | 0.048 (0.030 , 0.066) | 1.92E-07 | 0.01% |
| Lymphocyte percentage | 0.009 (-0.007 , 0.025) | 0.291 | 0.00% |
| Basophil percentage | -0.003 (-0.006 , 0.000) | 0.075 | 0.00% |
| Eosinophil percentage | -0.010 (-0.014 , -0.006) | 1.30E-05 | 0.01% |
| Monocyte percentage | -0.010 (-0.015 , -0.004) | 1.39E-03 | 0.00% |
| Ethnicity (ref: White) |  |  | 0.84% |
| Mixed | 0.126 (0.088 , 0.164) | 8.80E-11 |  |
| Asian | 0.049 (0.028 , 0.071) | 5.07E-06 |  |
| Black | 0.412 (0.387 , 0.436) | 1.41E-245 |  |
| Chinese | 0.359 (0.308 , 0.411) | 2.00E-42 |  |
| Other | 0.185 (0.155 , 0.216) | 2.21E-32 |  |

#### Variance in LTL explained

In a multivariable model, we estimated the amount of variance in LTL explained by the biological factors studied, excluding parental age at birth which was only available for a small fraction of the cohort. Age explained ∼3.5%, followed by ethnicity, sex and WBC, explaining 0.84%, 0.68% and 0.37%, respectively (**Table 6**). Allowing for WBC, blood cell proportions individually accounted for very little additional variance (all <0.01%, **Table 6**). In aggregate, these factors explained about 5.5% of the variance in LTL. In this model, where cell composition is also included, we also detected a significant difference in LTL between White participants and the category in UKB called “Asians” (comprising mostly South Asians). However, the difference in LTL was most marked for Black and Chinese ethnicities where the difference in “biological age” compared to White participants was 17.9 years and 15.6 years, respectively (**Table 6**).

## Discussion

We have generated relative LTL measurements in 474,074 well-characterised participants in UKB, creating an unprecedentedly powerful resource to investigate the determinants and biomedical consequences of naturally-occurring variation in LTL. Removing technical variation from the measurements through careful curation of relevant variables and statistical adjustment improved measures of inter-assay variation and led to a more stable measurement of LTL over the 4-year measurement period. Despite the unprecedented scale of the project, our assay showed good reproducibility as assessed through inclusion of both blinded as well as deliberate duplicates.

Our confirmation of well-established relationships between shorter LTL and older age and male sex of similar magnitudes to those reported before adds confidence to the validity of our measurements. For example, our estimate that women are younger in “biological age” than men by 7.4 years is very similar to an estimate of 7.0 years based on previous data^30^. Our study’s exceptional power allowed us to demonstrate a moderate but significant age-sex interaction in the inverse association of LTL with age, showing shallower associations in younger women compared with men but more similar associations after the menopause or after age 55 years. This observation is consistent with a potential protective effect of oestrogen on LTL attrition^31^. However, our analysis was constrained by the relatively narrow age at recruitment of participants in UKB (40-70 years); other studies have reported steeper associations of shorter LTL with age in younger women^32,33^. Furthermore, the cross-sectional design of both UKB and the other studies that have investigated sex-related associations of LTL with age, limit the inferences that can be drawn; longitudinal studies are needed to confirm any oestrogen-related associations with LTL.

Our study found that longer LTL is associated with having an older father at the time of birth, again consistent with previous findings^7,10,11^. Although we also observed an association between longer LTL and having an older mother at birth, additional analysis showed that this was most likely due to correlation of spousal ages and the association is driven predominantly, if not exclusively, through paternal age at birth. It is notable, therefore, that previous studies have reported longer telomeres in the sperm of older men^10^.

We also observed substantial ethnic differences in average LTL, confirming previous findings of longer LTL in people of African ancestry^12-14^. Furthermore, compared to people of white European ancestry, we report novel findings of longer LTL in people of Chinese, South and West Asian and mixed ancestry. The reasons for these differences and any potential biomedical consequences remain to be explored.

There has been a long debate about the potential impact of white cell composition on LTL measurements prompted by previous reports of differences in TL between B cells, T cells and monocytes within an individual^34-37^. Here we clarify that, at a population level, total white cell count has a small but significant inverse association with LTL. Accounting for this, the proportions of several white cell types available in UKB additionally explained very little of the population variance in LTL.

Using paired samples from 1351 participants taken on average 5 years apart, we show the regression-dilution ratio for LTL is ∼0.65. This degree of within-individual variability is similar to those we observed for systolic blood pressure and total cholesterol, but less than for body mass index in the same UKB participants (**Supplementary Table 1**). A previous study, involving a larger number of paired measurements, reported a somewhat lower regression-dilution ratio (∼0.50) for LTL, perhaps because the interval between measurements was more prolonged (9.3 vs 5.5 years), meaning age-related changes in LTL could have contributed more substantially. The implication from both of these studies is that, despite its high heritability, LTL is a fluctuating factor within individuals in mid-life. Hence, adjusting for RDR should provide a more accurate assessment of any aetiological associations of LTL with disease outcomes and biomedical traits.

As noted earlier, UKB combines several key attributes that make it an exceptionally informative cohort in which to conduct LTL measurements. UKB is not, however, a strictly representative sample of the UK general population, as only about 6% of those invited to participate did so^38^. Risk factor levels and mortality rates in UKB are lower than in the general population due to a “healthy cohort” effect. Nevertheless, LTL-phenotype associations from UKB should have strong internal validity and, given the diverse characteristics of this large cohort, should be applicable to many different settings^39^.

In summary, we present a large, high-quality resource to facilitate investigation of the determinants and biomedical consequences of inter-individual variation in LTL. We provide a detailed description of generation and quality assurance of the measurements. Demonstration of several well-established relationships of LTL should give researchers additional confidence in the use of the resource.

## Methods

### Measurement of LTL

Technicians at UKB extracted DNA from peripheral blood leukocytes as part of a cohort-wide array genotyping project, described in detail elsewhere^40^. DNA was extracted using an automated process for the majority of samples; a small proportion were extracted using a manual method using the same chemistry. UKB transported residual DNA from this project to the University of Leicester LTL assay laboratory in 11 tranches of approximately 50,000 samples. Sample manifests including sample ID and concentration were provided alongside the samples. Prior to assay, samples were first normalised to a concentration of 10ng/ul using automated pipetting robots (Qiagility, Qiagen). Research staff at the University of Leicester conducted LTL measurements blinded to phenotypic information.

Using the multiplex qPCR methodology LTL is measured as the ratio of telomere repeat copy number (T) relative to that of a single copy gene (S, Hgb)^41^. The amounts of both T and S were measured within each reaction and were calculated relative to a calibrator sample (pooled DNA from 20 individuals) which was included on every run. Each measurement run was set up on a 100 well Rotor-Disc (Qiagen) using an automated pipetting robot (Qiagility, Qiagen) and included 47 samples in duplicate, a no template control and the calibrator sample in quadruplicate. Each qPCR reaction contained 1x Sensimix SYBR No-ROX enzyme mix (Bioline), 150nM Tel primers, 45nM of Hgb primers (**Supplementary Table 2**) and 30ng of DNA. The Rotor-Discs were transferred to a Rotor-Gene Q PCR machine for amplification. Cycling conditions for each run were as follows: 95°C 10 min; 95°C for 15 sec, 49°C for 15 sec for 2 cycles; 94°C for 15 sec, 62°C for 10 sec, 72°C for 15 sec with signal acquisition (T), 84°C for 10 sec, 88°C for 10 sec with signal acquisition (S) for 32 cycles. At the end of cycling a dissociation curve was included. Prior to use, each primer batch was assessed for quality by producing a standard curve across the input DNA range of 1200-9.3ng in two-fold dilution (8 points). Primers achieving 90-110% reaction efficiency and an R^2^ across the linear range >0.99 were acceptable. Further testing was then performed to reproduce measurements for previously assayed samples with good concordance before further use. The linear range for each primer batch was recorded as a QC metric.

Relative quantities of T and S were calculated for each sample using the Rotor-Gene comparative quantification software (Qiagen). This software calculates the amplification efficiency of each reaction. The relative amount of T and S is calculated using the following equation:

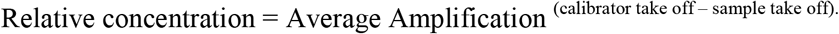

Using the calculated average amplification efficiency, rather than assuming 100% efficiency, effectively adjusts the measurements for run-to-run variation. The resulting T/S ratios were calculated for each well, alongside the average T/S and the coefficient of variation for the sample duplicate. We then applied strict, pre-defined QC criteria at both the sample and run levels, as detailed in **Supplementary Table 3**, before accepting the measurements as being valid. Following this, successful data from each run was uploaded into a custom database. All samples that failed QC criteria were re-assayed until valid measurements were achieved, or the sample was deemed to be unsatisfactory or exhausted.

To measure stability and reproducibility of the measurements subsets of samples were deliberately re-run at later dates and the coefficient of variation between the measurements calculated. For this subsets of samples were selected each week and re-measured. These samples were deliberately selected from early tranches so that as the project progressed reproducibility could be assessed over longer time periods. In addition to these deliberate repeats (n=22,516), a small number of duplicate samples (n=528) were included by UKB, spread across the tranches, to which we were initially blinded (blinded duplicates).

Due to the scale of the project, the samples were measured over a 47 month period by 6 members of staff (“operators”), using 5 Qiagility pipetting robots for liquid dispensing and 8 Rotor-Gene PCR machines (**Supplementary Figure 11**). It was necessary to use 19 batches of Sensimix SYBR No-ROX enzyme mix and 7 primer batches for the assays (**Supplementary Figure 12**). Details of these parameters, alongside temperature and humidity (for potential influences on Rotor-Gene and Qiagility performance), were recorded alongside the sample data.

### Statistical adjustment of data to minimise technical variation

Adjustment for T/S experimental/technical variation was performed in three stages using R v3.6.1. First, backwards selection using the mean T/S ratios at the run level was used in a linear regression adjusting for enzyme, primer batch, PCR machine, pipetting robot, operator, temperature, humidity, time of day, and extraction method. Only runs with at least 20 valid measurements were included. Significant effects were determined using the Bayesian information criterion. The second stage took all significant main effects identified in stage 1 and further tested all possible two-way interactions using the same backwards selection approach as stage 1 for the interaction effects. For both stages we estimate a partial R^2^ as the difference between the full model R^2^ and the model R^2^ leaving a single parameter out. Individual-level T/S ratios were then partially adjusted based on the coefficients from the final model selected in stage 2. A further level of adjustment was then applied by fitting a linear regression model on the individual level data adjusting for the 260/280 ratio of the DNA sample (stage 3). Due to an observed non-linear relationship between the T/S and 260/280 ratios both linear and quadratic effects were included. For the purpose of this analysis samples with a missing 260/280 or those that had a measurement within the extremes of the distribution (<1 or >3) were imputed using the mean 260/280 value.

After technical adjustments were applied the LTL measurements (T/S ratios) were log_e_-transformed due to non-normality (log_e_-LTL). To allow direct comparison of the results of our analyses with previous studies we Z-standardised the log_e_-LTL measures.

### Estimation of regression dilution bias

DNA was extracted by UKB for 1884 participants from a second blood sample taken between 2 and 10 years after the original sample, using the same methodology. To remove technical variation between the two measures for estimation of the regression-dilution the original baseline sample was re-plated alongside the second time point sample and 23 pairs of samples were assayed in each qPCR run. As these DNA samples were received towards the end of the project, for many there was insufficient DNA remaining from the baseline sample (which had already undergone measurement) to allow measurements for both of the paired DNAs to be obtained. Quality control parameters were then applied as for the main dataset. Only samples with valid data for both time points within the same run were taken forward for analysis (**Supplementary Figure 13**).

We estimated the LTL regression dilution ratio (RDR) coefficient by regressing LTL measured at the second time point on LTL measured at the first time point^27^. The RDR is the ratio of the between-individual variance to the total variance (i.e. between-individual variance + within-individual variance); RDR values close to 1 indicate little within-individual variability, whereas values close to 0 imply high levels of within-individual variability. The resulting regression coefficient is the RDR, and the multiplicative regression dilution bias (RDB) correction factor, λ, is simply the inverse of the RDR coefficient i.e.

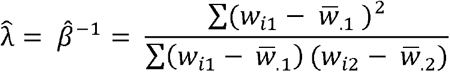

Where *w*_*i*1_ and *w*_*i*2_ are the first and second measurements of LTL respectively for each of the 1,351 participants.

We further adjusted for the difference in ages between the two measurements to consider the impact of time between sample collections on the RDR estimate and after removing the age effect from the first and second measurements by taking the residuals from a linear regression on LTL adjusted for age. We then regressed the age-adjusted second measurement residuals on the age-adjusted first measurement residuals adjusting for baseline age, sex and difference in age between sample collections to estimate the RDR. For non-LTL traits in UKB shown in **Supplementary Table 1**, we used baseline and follow-up visit 1 data and ran the models in the same way to estimate the RDR.

### Association of LTL with selected phenotypes in UKB

Before conducting analyses we first removed participants for whom the LTL measurement was made from a non-baseline sample (where baseline visit date was before sample collection date) or where self-reported sex and genetic sex did not match (reflecting potential sample mishandling)^29^. To assess population demographics we estimated means and standard deviations for continuous traits and percentages for categorical traits. To account for familial correlation we randomly excluded one participant from each related pair, where a pair of participants were related if their kinship coefficient was K>0.088 estimated using genetic relatedness. We used linear regression models to assess the association of TL with age, sex, parental age at birth, ethnicity and white blood cell traits. Interactions and non-linear effects were considered in the regression model where appropriate. We consider P<0.05 as the threshold for nominal statistical significance.

Age and sex relationships were assessed first to identify interactions and non-linear effects in the data to estimate population attrition rates. To further investigate the observed age and sex trends we investigated the role of menopause by matching a male to each female 1:1 on age at baseline running stratified analyses by pre- and post-menopause status.

We calculated parental age at birth from the reported parental age at baseline minus the age of the participant at baseline. We first modelled parental age at birth adjusting for age and sex and then calculated the difference in paternal and maternal age running analyses stratified by age difference group, 2-5 years and >5 years and run separately. Similarly, for ethnicity, regression models were stratified by ethnic group and run separately to assess the age and sex attrition rates within each ethnic group. We used the UKB defined ethnic groups from self-reported data (Data-Field 21000). Both “British and Black British” and “Asian and British Asian” are shortened to “Black” and “Asian” throughout. The “Asian and British Asian” is largely comprised of South and West Asian ancestries. We considered collinearity in these models through estimation of the variance inflation factor (VIF) where a value > 5 is considered to indicate collinearity.

Finally, we fit a multivariable model to assess the contribution of white blood cell traits. All white blood cell traits were winsorized at the 0.5% and 99.5% centile to reduce the impact of extreme values, log-transformed if required and Z-standardised. Linear regression models were again used to quantify the association with total white blood cell count on TL. We also included white blood cell composition in the model with the percentages of neutrophils, monocytes, eosinophils, lymphocytes and basophils. All phenotype analyses were run using Stata v16.0.

## Data Availability

Data is available to researchers through application to UK Biobank

## Acknowledgements

This research has been conducted using the UK Biobank Resource under Application Number 6077 and was funded by the UK Medical Research Council (MRC), Biotechnology and Biological Sciences Research Council and British Heart Foundation (BHF) through MRC grant MR/M012816/1. C.P.N is funded by the BHF (SP/16/4/32697). V.C., C.A.B., C.M., V.B., Q.W., R.B., C.P.N. and N.J.S. are supported by the National Institute for Health Research (NIHR) Leicester Cardiovascular Biomedical Research Centre (BRC-1215-20010). Cambridge University investigators are supported by the BHF (RG/13/13/30194; RG/18/13/33946), Health Data Research UK, NIHR Cambridge BRC (BRC-1215-20014), NIHR Blood and Transplant Research Unit in Donor Health and Genomics (NIHR BTRU-2014-10024) and MRC (MR/L003120/1). J.N.D. holds a BHF Personal Professorship and NIHR Senior Investigator Award. A.M.W. and E.A. received support from the EU/EFPIA Innovative Medicines Initiative Joint Undertaking BigData@Heart grant n° 116074. Parsa Akbari, Thomas Bolton and Matthew Arnold made computational and biostatistical contributions to this work. We thank Louise Courtney, Samantha Welsh and Daniel Fry for assistance with UK Biobank samples.

## Author contributions

M.D., C.S, M.P., S.Sh., D.E.N. and V.C. generated the data. S.C.W., C.A.B., R.B., J.R.T., V.C. and C.P.N. curated the data. C.M., V.B., Q.W., A.S.B., J.R.T., V.C. and C.P.N performed statistical analyses. V.C., C.P.N., C.M., Q.W., C.A.B., E.A., S.K., S.St., V.B., T.J., E.D.A., A.M.W., A.S.B., J.R.T., J.N.D. and N.J.S. drafted the manuscript and all authors revised it. V.C., C.P.N., J.R.T., J.N.D. and N.J.S. (Principal investigator) secured funding and oversaw the project.

