## Supplementary Tables and Figures for "A major population resource of 474,074 participants in UK Biobank to investigate determinants and biomedical consequences of leukocyte telomere length"

Supplementary Figures and Tables

**Contents**

Supplementary Figures	2

Supplementary Tables	15

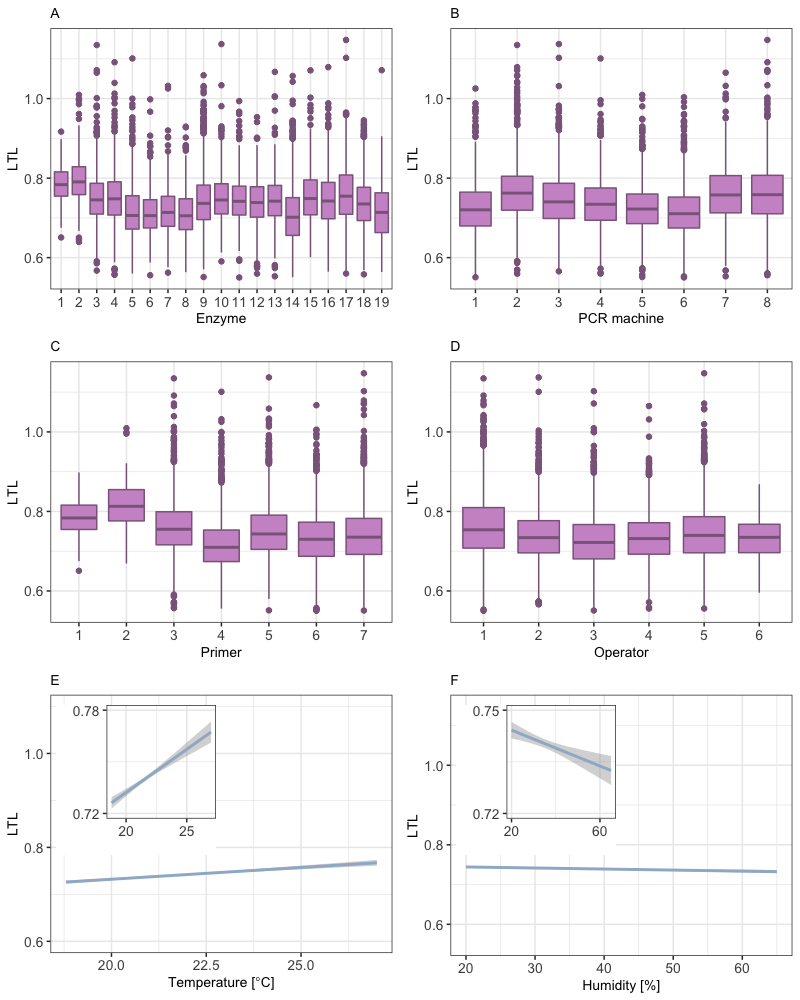

**Supplementary Figure 1. Significant technical parameters affecting LTL measurements based on the stage 1 adjustment.** Summary boxplots are shown for LTL measurements for each associated parameter: Enzyme batch (A), PCR machine (B), primer batch (C), operator (D). Linear relationships were seen between LTL and temperature (E) and humidity (F).

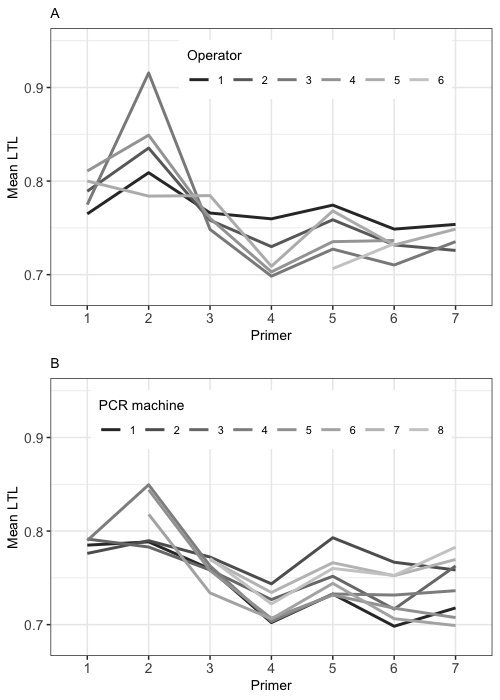

**Supplementary Figure 2. Significant interactions based on the stage 2 adjustment**. A) LTL by Primer and Operator. B) LTL by primer and PCR machine. PCR machines 5 and 6 were not used at the start of the pilot study (primer batch 1) and machines 7 and 8 were used from the end of the pilot stage (primer batch 3 onwards).

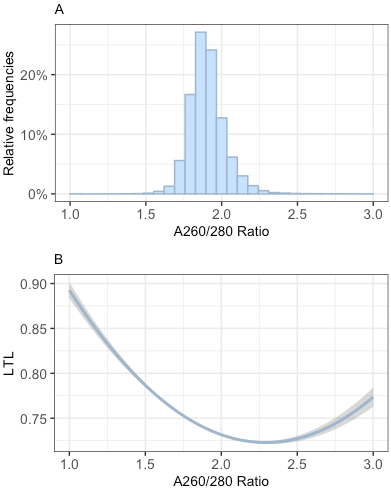

**Supplementary Figure 3. Effect of A260/280 on LTL.** The distribution of DNA sample A260/280 ratios is illustrated in (A). We observed an increase in LTL with very low and very high A260/280 ratios (B).

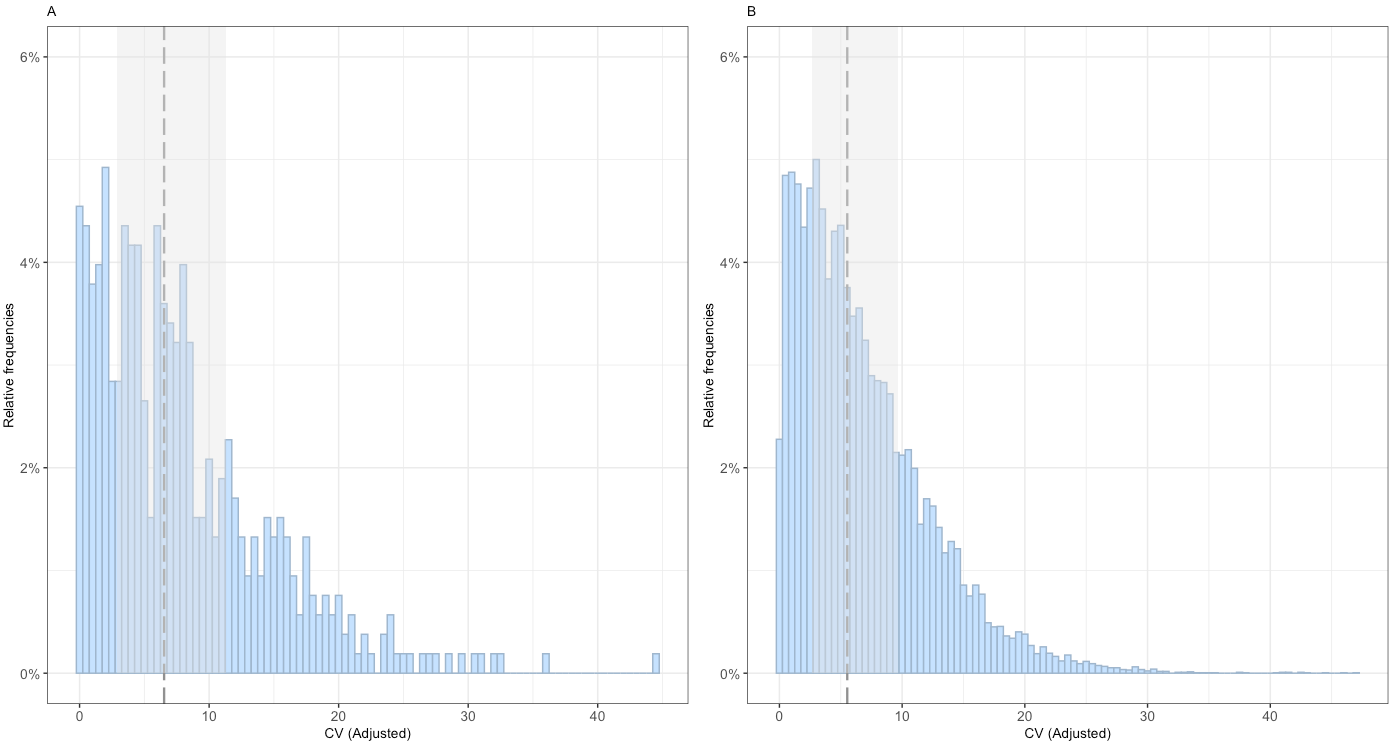

**Supplementary Figure 4: Distribution of the coefficients of variation for the repeat samples.** Distribution of CVs after technical adjustment for both the *blinded* repeats A) (n=528) and *deliberate* repeats B) (n=22,615) are shown. The grey dotted line represents the median coefficient of variation with the shaded region representing the interquartile range.

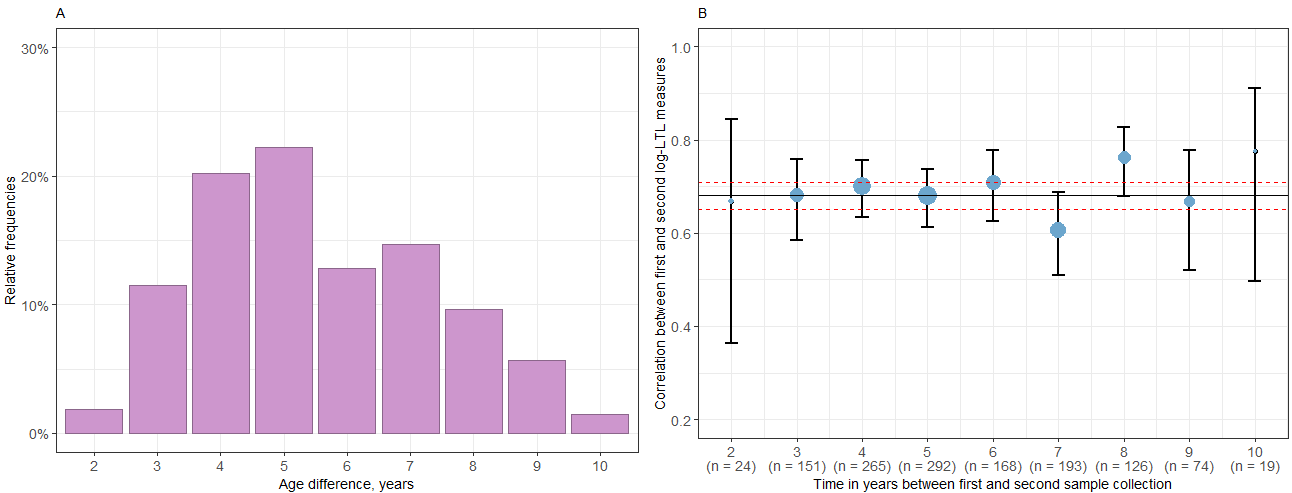

**Supplementary Figure 5.** **Data on the first and second DNA sample used to estimate regression dilution ratio.** A) Histogram showing that the gap between the two sample collections has a mean interval of 5.5 years (range: 2-10 years). B) Correlation between the first and second log-LTL measure by time, estimated as the difference in years between the two sample collections and shown with 95% confidence intervals. The size of the blue circle reflects the number of participants measured each year, shown in brackets. The black line shows the overall pooled correlation for all samples and the red dotted lines indicate the 95% confidence interval for this estimate.

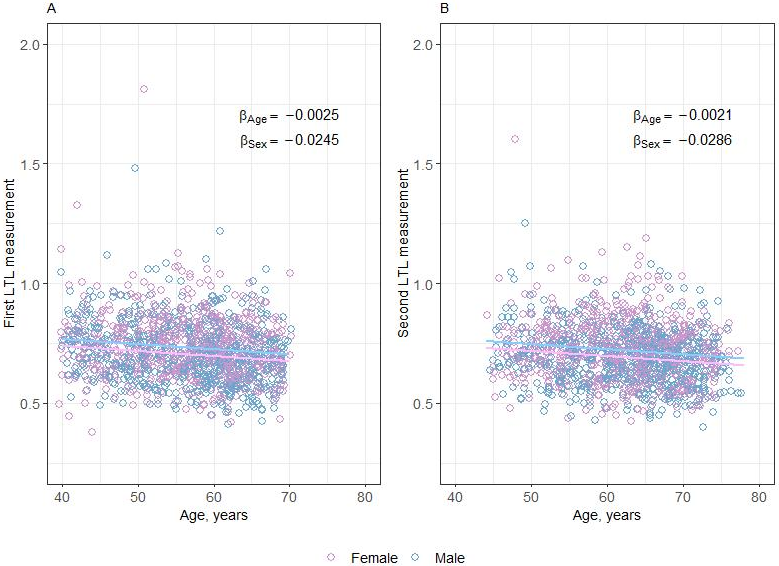

**Supplementary Figure 6**: **Age and sex relationships for participants used to estimate regression dilution bias.** The decline of LTL with age is shown for men (blue) and women (plum) for both the first (A) and second (B) LTL measurements. The estimated effect sizes are shown for age ($\beta_{Age}$) and sex ($\beta_{Sex}$) within the figures.

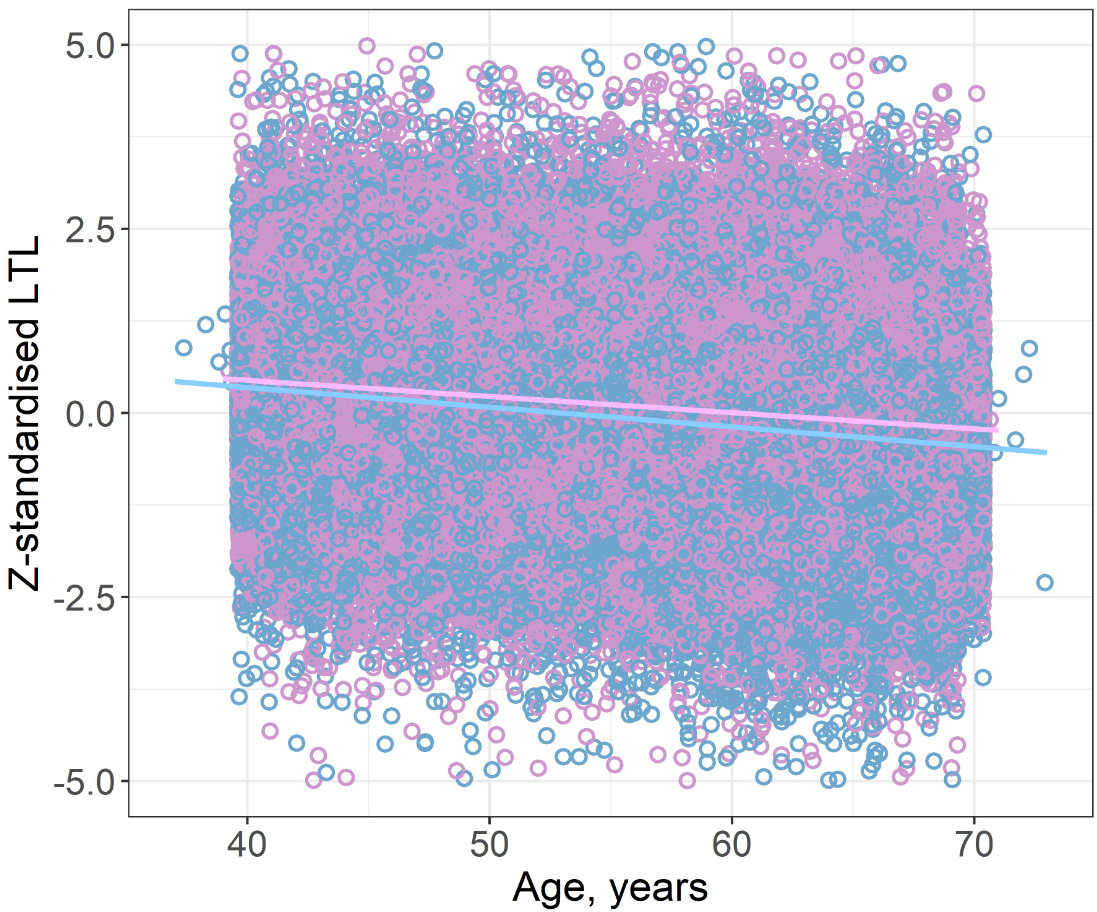

**Supplementary Figure 7 Decline of LTL with age.** The decline of z-standardized log_e_-LTL with age is shown for men (blue) and women (plum) in adjusted data. The y-axis is truncated at -5SD to +5SD with 166 data points (80 women, 86 men) not shown. A small number of participants recruited by UK Biobank fall outside of the stated 40-69 age range.

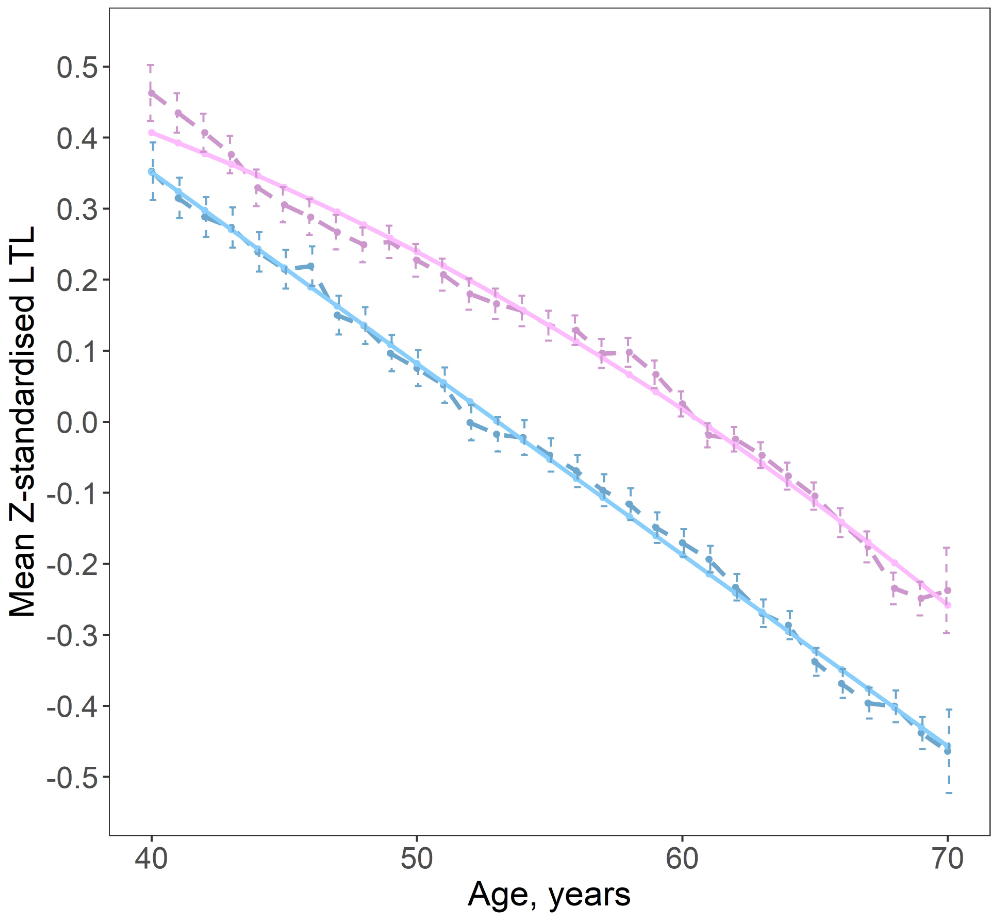

**Supplementary Figure 8. Decline in LTL with age by sex**. Using stratified regression for men (blue) and women (plum) we considered the non-linear effect of age within each sex. Here we show the predicted shape in a solid line and the observed data in a dashed line with confidence intervals. There is significant non-linearity observed for women, where the rate of LTL decline increases as the population ages.

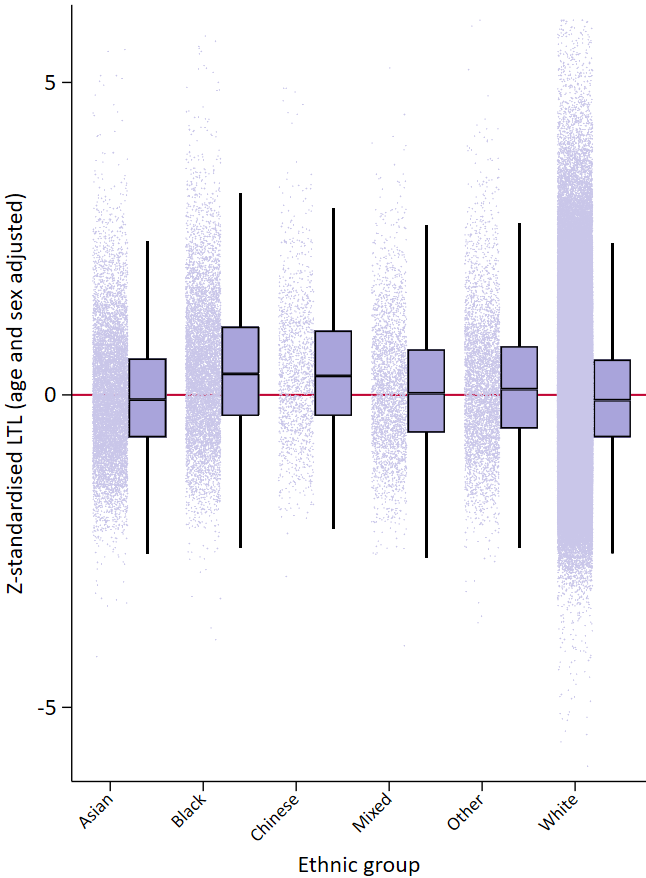
**Supplementary Figure 9. Box plot of telomere length within individual ethnic groups.** Data are adjusted for both age and sex with the median and interquartile range shown within the box plot. Individual observations also shown to indicate the range and quantity of data. Ethnicity is self-reported and presented as defined by UK Biobank Data-Field 21000. Note that we shorten “Asian or Asian British” to Asian and “Black or Black British” to Black.

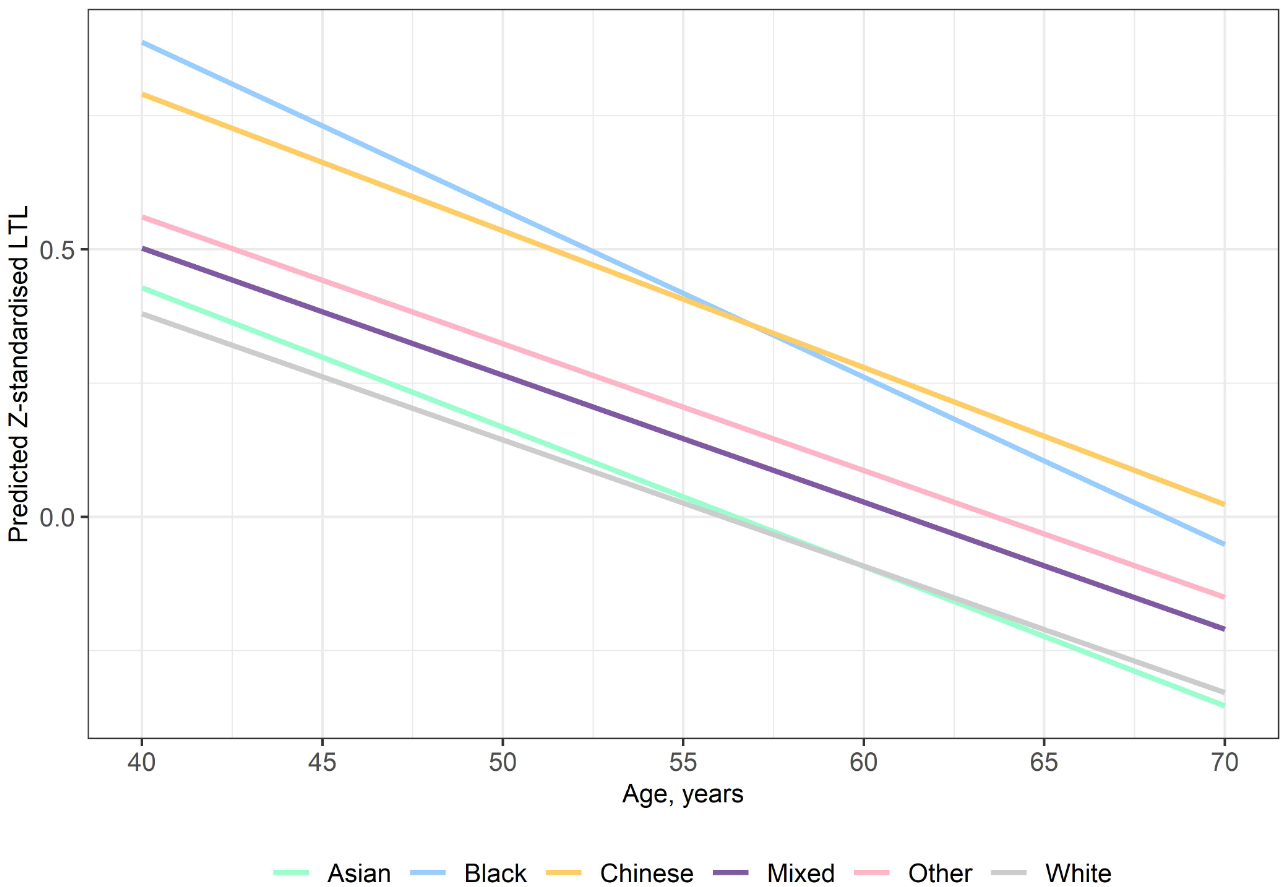

**Supplementary Figure 10. Sex adjusted LTL by age in different ethnic groups**. The rate of population decline in LTL with age is steeper in the Black population (blue line).

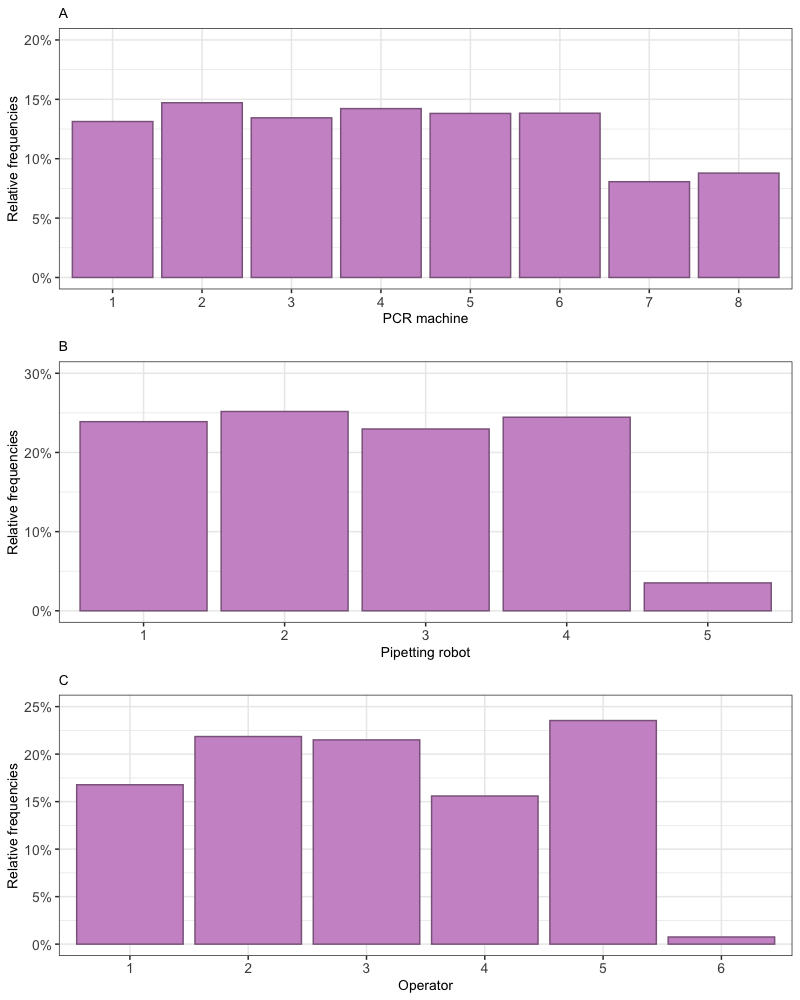

**Supplementary Figure 11. Relative frequencies of samples measured by operator or machine**. A) Eight PCR machines were used throughout the project. A higher proportion of samples were run on machines 1-6. B) Four pipetting robots were used predominantly throughout the project, with only a small proportion being run on the machine used to cover mechanical breakdown. C) Five main operators ran the majority of the samples.

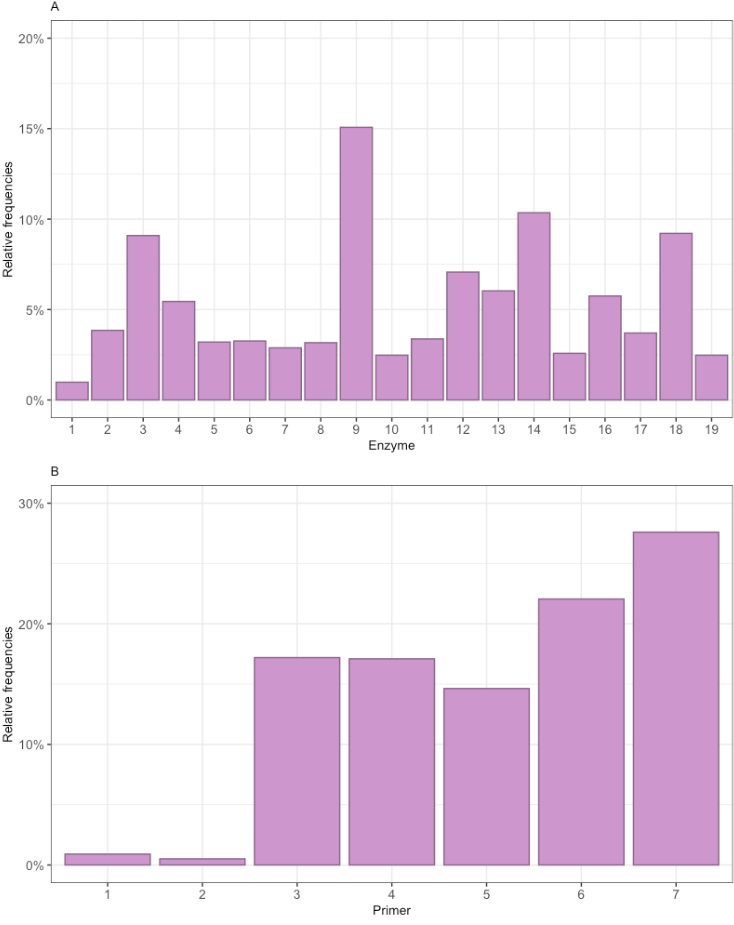

**Supplementary Figure 12. Relative frequencies of samples measured with different reagent batches**. A) The proportion of samples run with each batch of enzyme is shown. Enzyme batch was defined as being from the same manufacturer’s lot number. B) The first two primer batches were used during the pilot study. Larger batches were made after the pilot to minimize batch effects.

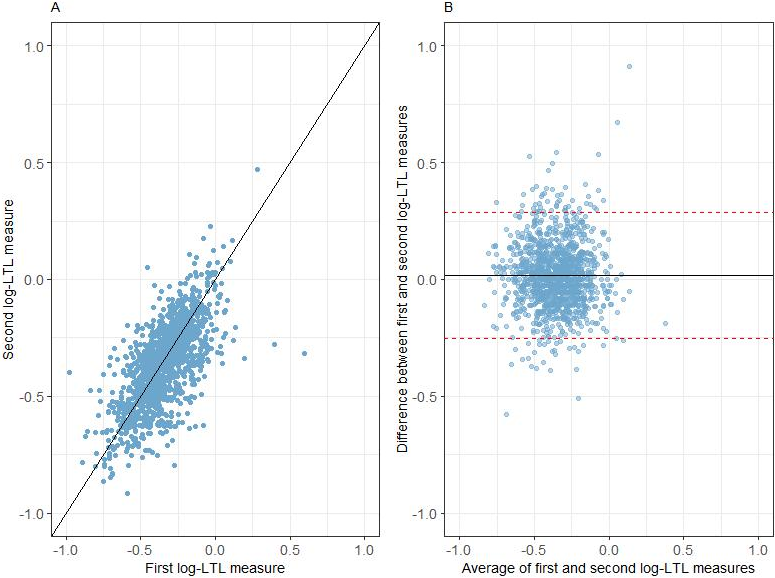

**Supplementary Figure 13**. **Correlation between first and second log-LTL measurement in the RDB experiment.** Data are shown for the 1,351 participants with valid data at both time points. The black line indicates perfect agreement. A) A scatter plot of the paired measurements. B) A Bland-Altman plot where the red dotted lines represent the 95% CI of the difference between the first and second LTL measure.

|  | **UK Biobank** | **Tehran lipid and glucose study** | **Emerging Risk Factors** | **4^th^ & 5^th^ Tromsø studies** |
| --- | --- | --- | --- | --- |
| **Participants in main study** | 502,540 | 6,327 | 302,430 | 5,970 |
| **Participants in RDB sub-study** | 20,346 | 3,063 | 89,073 | 5,179 |
| **Time between studies** | 4.4 years | 3 years | 4.9 years | 7 years |
| **Systolic blood pressure** |  |  |  |  |
| Men | 0.633 | 0.690 |  |  |
| Women | 0.719 | 0.741 |  |  |
| Overall | 0.680 |  | 0.521 | 0.625 |
| **Diastolic blood pressure** |  |  |  |  |
| Men | 0.592 | 0.654 |  |  |
| Women | 0.667 | 0.610 |  |  |
| Overall | 0.633 |  |  | 0.578 |
| **Body mass index** |  |  |  |  |
| Men | 0.943 | 0.926 |  |  |
| Women | 0.935 | 0.926 |  |  |
| Overall | 0.943 |  | 0.962 | 0.935 |
| **Triglycerides** |  |  |  |  |
| Overall | 0.571 | 0.685 | 0.629 | 0.478 |
| **Cholesterol** |  |  |  |  |
| Overall | 0.694 | 0.714 | 0.599 | 0.556 |
| **LDL** | 0.676 |  |  |  |
| **HDL** | 0.893 | 0.633 | 0.690 | 0.725 |
| **HbA1c** | 0.758 |  |  |  |

**Supplementary Table 1. Estimates of RDB for cardiovascular risk factors.** Data are taken from the available literature, except for UK Biobank data that were estimated in project 6077.

| Name | Sequence 5’ – 3’ | Purification |
| --- | --- | --- |
| Telg | ACACTAAGGTTTGGGTTTGGGTTTGGGTTTGGGTTAGTGT | HPLC |
| Telc | TGTTAGGTATCCCTATCCCTATCCCTATCCCTATCCCTAACA | HPLC |
| Hgbu | CGGCGGCGGGCGGCGCGGGCTGGGCGGCTTCATCCACGTTCACCTTG | PAGE |
| Hgbd | GCCCGGCCCGCCGCGCCCGTCCCGCCGGAGGAGAAGTCTGCCGTT | PAGE |

**Supplementary Table 2.** **Primer sequences and purification levels.**

|  | **Reason for QC failure** | **Criteria** |
| --- | --- | --- |
| Half plate level | Contamination | Amplification in NTC wells not attributable to primer dimer formation |
|  | Inaccurate calibration | Less than three valid measurements for calibrator |
|  |  | Calibrator sample CV≥10% |
|  |  | Extreme mean LTL across half plate |
|  | Incorrect master mix preparation | More than 5 samples with CV≥12% |
|  |  | Average amplification >0.2 from what is usual for the machine for either T or S |
|  |  | Standard deviation of amplification efficiency >0.2 after removal of failed samples and NTCs |
|  |  | Melt curve does not conform to the expected pattern for more than 5 samples |
|  | Inaccurate pipetting | More than 5 samples with CV≥12% |
| Sample level | Inaccurate pipetting | Sample CV≥10%* |
|  |  | Failure of one replicate to amplify |
|  | Potential sample quality issues | Odd amplification (>1.5 from mean) |
|  |  | Melt curve does not conform to the expected pattern |
|  | Insufficient DNA quantity/inaccurate sample dilution | Failure of one replicate to amplify |
|  |  | Amplification outside linear range of the assay |

**Supplementary Table 3. Details of QC assessment parameters applied at the half plate and individual sample level.** Likely technical reasons contributing to poor data quality are shown alongside the relevant criteria to signify a QC failure. It should be noted that certain criteria are not exclusively attributable to a single technical problem. For instance, a high number of samples with a CV≥12% could indicate either incorrect primer concentrations within the master mix or inaccurate dispensing of the master mix and/or samples into the reaction plate. Samples failed QC if samples met any of the listed criteria. *An intra-sample CV>10% was a pre-set criterion to try and ensure we obtain the most reliable data. It should be noted that criteria are not mutually exclusive.
